# Incidence, prevalence, and survival of lung cancer in the United Kingdom from 2000-2021: a population-based cohort study

**DOI:** 10.1101/2024.02.26.24303406

**Authors:** George Corby, Nicola L Barclay, Eng Hooi Tan, Edward Burn, Antonella Delmestri, Talita Duarte-Salles, Asieh Golozar, Wai Yi Man, Ilona Tietzova, OPTIMA Consortium, Daniel Prieto-Alhambra, Danielle Newby

**Affiliations:** Centre for Statistics in Medicine, Nuffield Department of Orthopaedics, Rheumatology and Musculoskeletal Sciences, University of Oxford, Oxford, OX3 7LD, UK; Odysseus Data Service, Cambridge, MA USA; OHDSI Center at the Roux Institute, Northeastern University, Boston, MA USA; Fundació Institut Universitari per a la recerca a l’Atenció Primària de Salut Jordi Gol i Gurina (IDIAPJGol), Barcelona, Spain; Department of Medical Informatics, Erasmus University Medical Centre, Rotterdam, The Netherlands; First Department of Tuberculosis and Respiratory Diseases, First Faculty of Medicine, Charles University, Prague, Czech Republic

**Keywords:** lung cancer, incidence, prevalence, cancer survival

## Abstract

**Background:** Lung cancer is the leading cause of cancer-associated mortality worldwide. In the UK, there has been a major reduction in smoking, the leading risk factor for lung cancer, as well as the introduction of the new screening in 2023. Therefore, an up-to-date assessment of the trends of lung cancer is required in the UK.

**Methods:** We performed a population-based cohort study using the UK primary care Clinical Practice Research Datalink (CPRD) GOLD database, compared with CPRD Aurum. Participants aged 18+ years, with one-year of prior data availability, were included. We estimated lung cancer incidence rates (IR), period prevalence (PP), and survival at one-, five– and ten years after diagnosis using the Kaplan-Meier method.

**Results:** Overall, 11,388,117 participants, with 45,563 lung cancer cases were studied. The incidence rate of lung cancer was 52.0 (95% CI 51.5 to 52.5) per 100,000 person-years, with incidence increasing from 2000 to 2021, especially in females aged over 50, and males aged over 80, with the highest incidence rate in people aged 80-89. Period prevalence in 2021 was 0.18%, with the largest rise seen in participants aged over 60. Median survival post-diagnosis increased from 6.6 months in those diagnosed between 2000-2004 to 10 months between 2015-2019. Both short and long-term survival was higher in younger cohorts, with 82.7% one-year survival in those aged 18-29, versus 24.2% in the age 90+ cohort. Throughout the study period, survival was longer in females, with a larger increase in survival over time than in males.

**Conclusion:** The incidence and prevalence of lung cancer diagnoses in the UK have increased, especially in female and older populations, with a small increase in median survival. With the introduction of the UK lung cancer screening programme, this study will enable future comparisons of overall disease burden, so the overall impact may be seen.

## 1. INTRODUCTION

Lung cancer is the leading cause of cancer-associated mortality worldwide, with 1.8 million deaths in 2020[1]. New diagnoses are predicted to nearly double by 2070 meaning it will continue to be a major cause of morbidity and mortality globally[2].

As early as 1962, the British Royal College of Physicians (RCP) published *Smoking and Health*[3], which established a clear and important link between smoking and lung cancer. Although smoking remains the greatest risk factor for all lung cancer subtypes, other factors such as family history[4], and exposure to arsenic[5], radon[6], biomass fuels, asbestos[7], or a broad range of occupational chemicals, have been identified as increasing risk. These risk factors can influence the proportion of patients with each subtype of lung cancer[2].

The UK Office of National Statistics (ONS) surveys indicate that 60 years from the initial RCP report, smoking prevalence in the UK continues to decrease, from 45% in the early 1970s to 14% in 2020[8], with a concurrent rise in the use of electronic cigarettes[9], [10]. This reduction in the prevalence of smoking has coincided with a revolution in treatments of lung cancer, with major advancements in surgery[11], targeted drug treatments[12], and a shift towards multidisciplinary team (MDT) management[13]. Together, this has corresponded with a major fall in mortality rates of lung cancer, with a 38% fall in mortality rate per 100,000 from the mid-1980s to late-2010s[13]. This reduction is driven by a decrease in male mortality, while female mortality has slightly increased from 1970, peaking in 2010, despite the prevalence of smoking falling in both sexes[13].

In 2023, the UK introduced targeted screening for lung cancer. People aged 55-74 with a GP health record documenting a smoking history will be invited to interview for a risk assessment, after which they may be offered a low-dose CT (Computed Tomography) scan. Focussing on those with the highest risk will enable earlier diagnosis and potentially better survival, with reduced iatrogenic harm from screening-related radiation exposure[14], [15].

Due to changes in risk factor exposure, and the introduction of the new screening in 2023, a comprehensive assessment of the trends of lung cancer in different population strata using routinely collected data from primary care, is required in the UK. Understanding these trends in lung cancer is an important aspect of population healthcare planning, particularly considering the introduction of screening high-risk individuals. Therefore, the aim of this study is to describe lung cancer burden and trends in terms of incidence, prevalence, and survival from 2000-2021 using two large and representative primary care databases from the UK.

## 2. METHODS

### 2.1 Study design, setting, and data sources

We carried out a population cohort study using routinely collected primary care data from the United Kingdom (UK). People with a diagnosis of lung cancer and a background cohort (denominator population) were identified from Clinical Practice Research Datalink (CPRD) GOLD to estimate overall survival, incidence, and prevalence. We additionally carried out this study using CPRD Aurum to compare the results for GOLD. Both these databases contain pseudonymised patient-level information on demographics, lifestyle data, clinical diagnoses, prescriptions, and preventive care provided to patients and collected by the NHS (National Health Service) as part of their care and support. CPRD GOLD contains data from across the UK whereas Aurum only contains data from England. Both databases are established primary care databases covering over 50 million people[16], and both were mapped to the Observational Medical Outcomes Partnership (OMOP) Common Data Model (CDM)[17], [18]. The use of CPRD data was approved by the Independent Scientific Advisory Committee (22_001843).

### 2.2 Study participants and time at risk

All participants were required to be aged 18 years or older and have at least one-year of prior history. For the incidence, prevalence, and survival analyses, the study cohort consisted of individuals present in the database from 1st January 2000. For CPRD GOLD, these individuals were followed up to whichever came first: practice stopped contributing to the database, patient left the practice, date of death, or the 31st of December 2021 (the end of the study period) whereas for Aurum, the end of the study period was 31st of December 2019. For the survival analysis, only individuals with newly diagnosed lung cancer were included. Any patients whose death and cancer diagnosis occurred on the same date were removed from the survival analysis.

### 2.3 Lung Cancer definition

We used Systematized Nomenclature of Medicine Clinical Terms (SNOMED CT) diagnostic codes to identify lung cancer events. Diagnostic codes indicative of either non-malignant cancer or metastasis were excluded as well as diagnosis codes indicative of melanoma and lymphoma occurring in the organs of interest. The study outcome cancer definition was reviewed with the aid of the CohortDiagnostics R package[19]. This package was used to identify additional codes of interest and to remove those highlighted as irrelevant based on feedback from clinicians with oncology, primary care, and real-world data expertise through an iterative process during the initial stages of analyses. The clinical code lists used to define lung cancer can be found in supplementary information S1.

OMOP-based computable phenotypes are available, together with all analytical code at our GitHub repository to enable reproducibility (see statistical methods). For survival analyses, mortality was defined as all-cause mortality based on CPRD GOLD date of death records, which have been previously validated and shown to be over 98% accurate[20].

### 2.4 Statistical methods

The population characteristics of patients with a diagnosis of lung cancer were summarised, with median and interquartile range (IQR) used for continuous variables, and counts and percentages used for categorical variables.

We calculated the overall and annualised crude incidence rates (IR) and annualised prevalence for lung cancer from 2000 to 2021. For incidence, the number of events, the observed time at risk, and the incidence rate per 100,000 person-years were summarised along with 95% confidence intervals (95% CI). Annual incidence rates were calculated as the number of incident lung cancer cases as the numerator and the number of person-years in the general population within that year as the denominator whereas overall incidence was calculated from 2000 to 2021.

Period prevalence was calculated on 1st January for the years 2000 to 2021, with the number of patients aged ≥ 18 years fulfilling the case definition for lung cancer as the numerator. The denominator was the number of patients ≥ 18 years on 1st January in the respective years for each database. The number of events, and prevalence (%) were summarised along with 95% confidence intervals.

For survival analysis, we used the Kaplan-Meier (KM) method to estimate the overall survival probability from observed survival times with 95% confidence intervals. We estimated the median survival and survival probability one, five, and ten-years after diagnosis.

All results were stratified by database, by age (ten-year age bands apart from the first and last age bands which were 18 to 29 years and 90 years and older respectively) and by sex. Additionally, for GOLD only, we stratified by calendar time of cancer diagnosis (2000-2004, 2005-2009, 2010-2014, 2015-2019 and 2020-2021) to understand if survival has changed over time. To avoid re-identification, we do not report results with fewer than five cases.

For Aurum, the same statistical analyses were performed using data from 1^st^ January 2000 to 31^st^ December 2019 to compare the results obtained from GOLD.

The statistical software R version 4.2.3 was used for analyses. For calculating incidence and prevalence, we used the IncidencePrevalence R package[21]. For survival analysis, we used the survival R package[22]. All analytic code used to perform the study is available at https://github.com/oxford-pharmacoepi/EHDENCancerIncidencePrevalence

## 3. RESULTS

### 3.1 Patient Populations and characteristics

Overall, there were 11,388,117 eligible patients, with at least one year of prior history identified from January 2000 to December 2021 from CPRD GOLD. Attrition tables for this study can be found in the supplement S2. A summary of baseline patient characteristics of those with a diagnosis of lung cancer is shown in Table 1.

**Table 1:** Baseline characteristics of lung cancer patients at the time of diagnosis for CPRD GOLD.

| Database | CPRD GOLD |
| --- | --- |
| Number of lung cancer patients | 45,563 |
| Sex: Male (N [%]) | 24569 (53.9%) |
| Age (years) (Median [IQR]) | 72 (65 to 79) |
| Age Groups N (years) (%) |  |
| 18-29 | 24 (0.1%) |
| 30-39 | 129 (0.3%) |
| 40-49 | 1030 (2.3%) |
| 50-59 | 4660 (10.2%) |
| 60-69 | 12249 (26.9%) |
| 70-79 | 16745 (36.8%) |
| 80-89 | 9546 (21.0%) |
| 90+ | 1180 (2.6%) |
| Prior history, days |  |
| median [IQR] | 3660 (2,009 to 5,352) |
| Smoking Status (any time five years prior) |  |
| Non-smoker | 7154 (15.7%) |
| Former smoker | 543 (1.2%) |
| Current smoker | 22019 (48.3%) |
| Missing | 15847 (34.8%) |
| General conditions (any time prior) |  |
| Atrial fibrillation | 3207 (7.0%) |
| Cerebrovascular disease | 3840 (8.4%) |
| Chronic liver disease | 247 (0.5%) |
| Chronic obstructive lung disease | 11163 (24.5%) |
| Coronary arteriosclerosis | 677 (1.5%) |
| Crohn's disease | 154 (0.3%) |
| Dementia | 772 (1.7%) |
| Depressive disorder | 6397 (14.0%) |
| Diabetes mellitus | 5321 (11.7%) |
| Gastroesophageal reflux disease | 1261 (2.8%) |
| Gastrointestinal haemorrhage | 3113 (6.8%) |
| Heart disease | 10704 (23.5%) |
| Heart failure | 2006 (4.4%) |
| HIV | 23 (0.0%) |
| Hyperlipidemia | 4586 (10.1%) |
| Hypertensive disorder | 12404 (27.2%) |
| Ischemic heart disease | 6237 (13.7%) |
| Obesity | 928 (2.0%) |
| Osteoarthritis | 9841 (21.6%) |
| Peripheral vascular disease | 2260 (5.0%) |
| Pneumonia | 2549 (5.6%) |
| Pulmonary embolism | 804 (1.8%) |
| Renal impairment | 6156 (13.5%) |
| Ulcerative colitis | 181 (0.4%) |
| Venous thrombosis | 2331 (5.1%) |

IQR: interquartile range

Overall, from the 45,563 patients with a diagnosis of lung cancer, patients were more likely to be male (54%), with a median age of 72 years old. The highest percentage of patients were aged 70-79 years old, contributing to 36.7% of diagnosed patients. Patients with lung cancer had a high proportion of COPD (25%) as well as cardiovascular comorbidities such as heart disease (23.5%) and hypertensive disorder (27.2%). Similar observations were seen across both databases. A similar table with detailed baseline characteristics for Aurum patients is available in Supplement S3.

### 3.2 Incidence rates stratified by calendar year, age, and sex

The overall IR of lung cancer from 2000 to 2021 was 52.0 (95% 51.5 to 52.5) per 100,000 person-years for GOLD. Females had lower overall IR (47.2 per 100 000 person-years (95% 46.5 to 47.8)) compared to males (57.0 per 100,000 person-years (95% 56.3 to 57.7)), with similar rates in Aurum. Annualised IRs increased across both databases (Figure 1). For GOLD, the annual IR dropped in 2020, before increasing in 2021. Females showed increasing IRs over the study period, while males showed a more stable trend in both databases. All study results can be found and downloaded in a user-friendly interactive web application: https://dpa-pde-oxford.shinyapps.io/LungCancerIncPrevSurvShiny/

**Figure 1:**
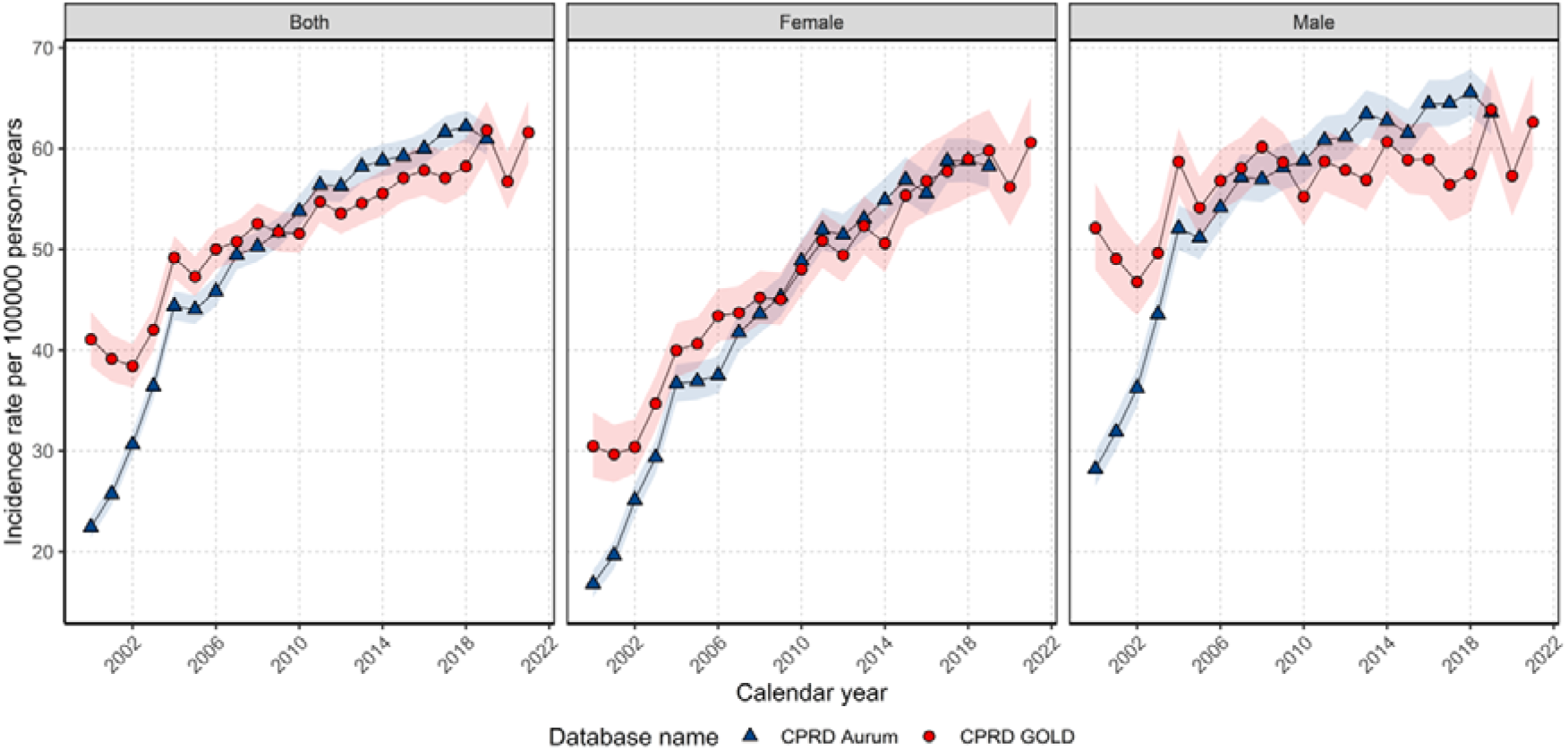
Annualised incidence rates for lung cancer from 2000 to 2021 stratified by database and sex. Bands show 95% confidence interval.

Overall IRs increased with older age up to 80–89 years. Those aged 18 to 29 years had the lowest overall IRs of 0.15 (0.10 to 0.22) per 100,000 person-years across the study period, whereas those aged 80–89 years had the highest IRs: 215.9 (211.6 to 220.2). Similar findings were obtained in Aurum (Supplement S4).

Annualised IRs for each age group (Figure 2) showed for those aged 70-89 a gradual increase between 2004-2019. Whereas for those 50-59 years of age there was a gradual decline in IRs since 2004. For those aged 60-69 years, IRs were stable from 2004-2019 for GOLD, whereas for Aurum, IRs increased from 2000 to 2013 before stabilising in 2019. For those over 90 years of age, there was also a stabilisation of IRs from 2014 with larger differences between the databases. Younger patients (30-49 years of age) showed relatively stable IRs over the study period.

**Figure 2:**
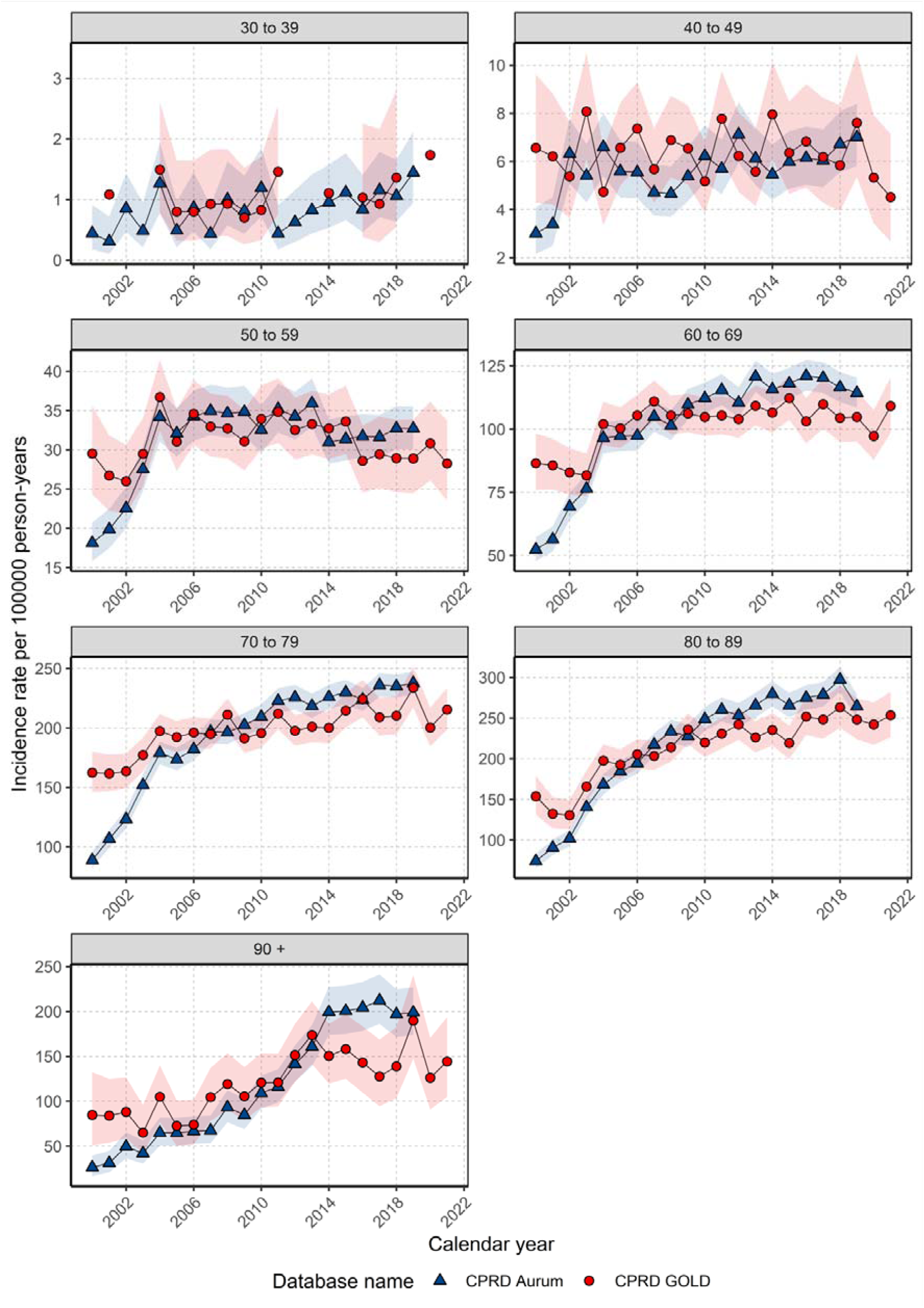
A**n**nualised **incidence rates from 2000 to 2021 stratified by database and age group. Bands show 95% confidence interval**.

Stratification by age and sex (Supplement S5) showed similar trends across both databases. For females aged 60-89 years, IRs increased between 2000-2019, whereas for males, IRs were either relatively stable across this time period. Males had higher IRs compared with females apart from those 30-59 years of age, where IRs were similar between sexes. Also, similar IRs were seen for females and males aged 60–69 years from 2015 onwards for GOLD (Supplement S5).

**3.3 Period prevalence for study population with database, age, and sex stratifications**

For the whole population, the PP for lung cancer in 2021 was 0.18% (0.17% to 0.18%) for GOLD. PP in 2019 was similar across both databases (∼0.17%). Sex stratification showed PP in 2019 were slightly higher in females (0.175% (95% 0.168% to 0.181%)) compared to males (0.156% (95% 0.150% to 0.162%)) in GOLD with smaller differences in Aurum. Trends in PP show a steep increase over the study period for the whole population and both sexes (Figure 3). In GOLD, PP has increased 2.7-fold from 2000 to 2021, with females having a larger fold increase (3.7-fold) in PP across the study period compared to males (2.0-fold), with similar observations seen in Aurum.

**Figure 3:**
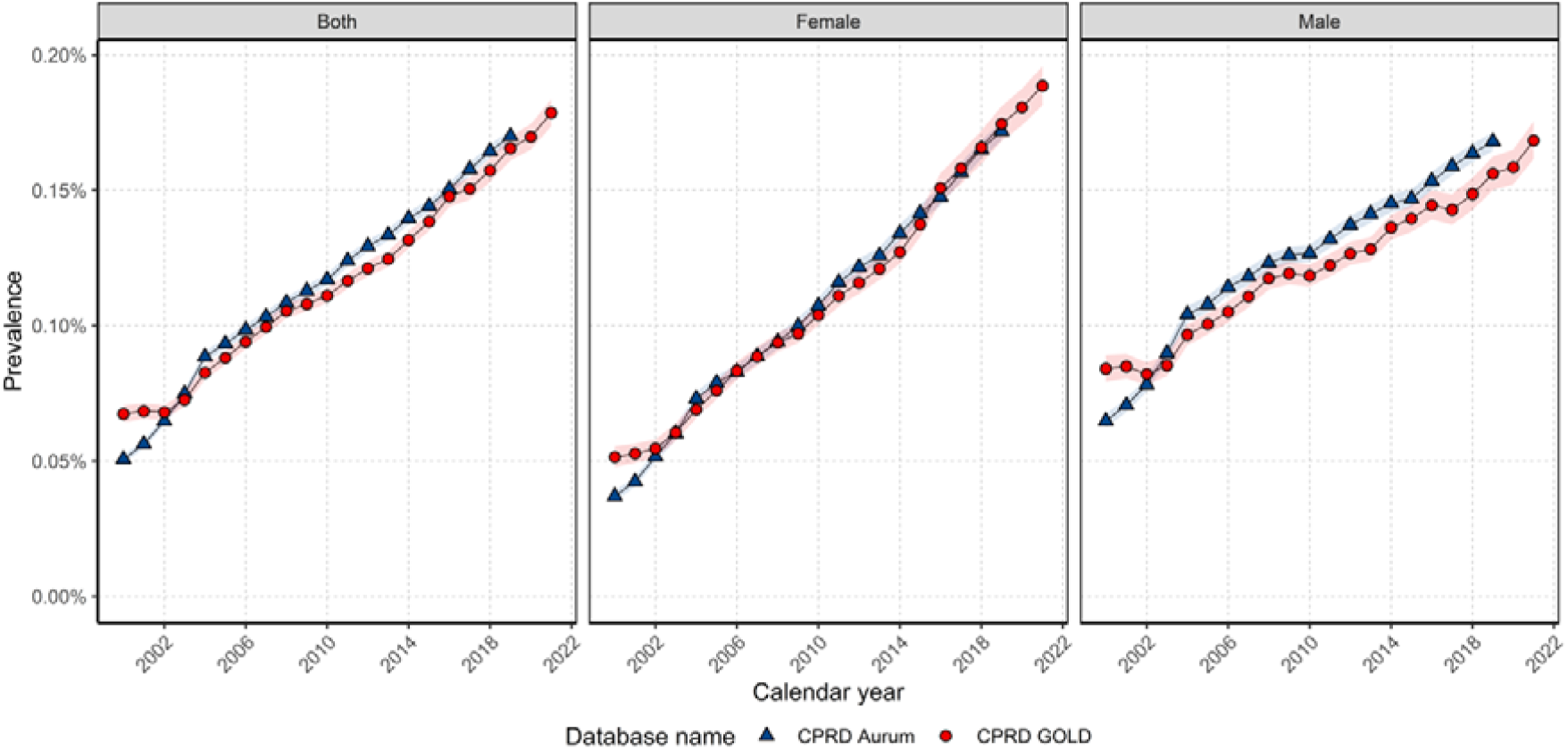
A**n**nual **period prevalence from 2000 to 2021 for the whole population and stratified by sex. Bands show 95% confidence interval**.

When stratifying by age group, PP in 2019 was highest in those 80-89 years of age (0.80% Aurum, 0.72% GOLD) with this age group seeing the largest change in PP over the study period (Figure 4). Overall, there were little differences in PP trends between age groups, with the majority of age groups showing increases in PP over the study period for both databases. Overall, PP increased from 2020 to 21 for GOLD for most age groups apart from those aged 40-49 years of age, for which increases in PP were minimal.

**Figure 4:**
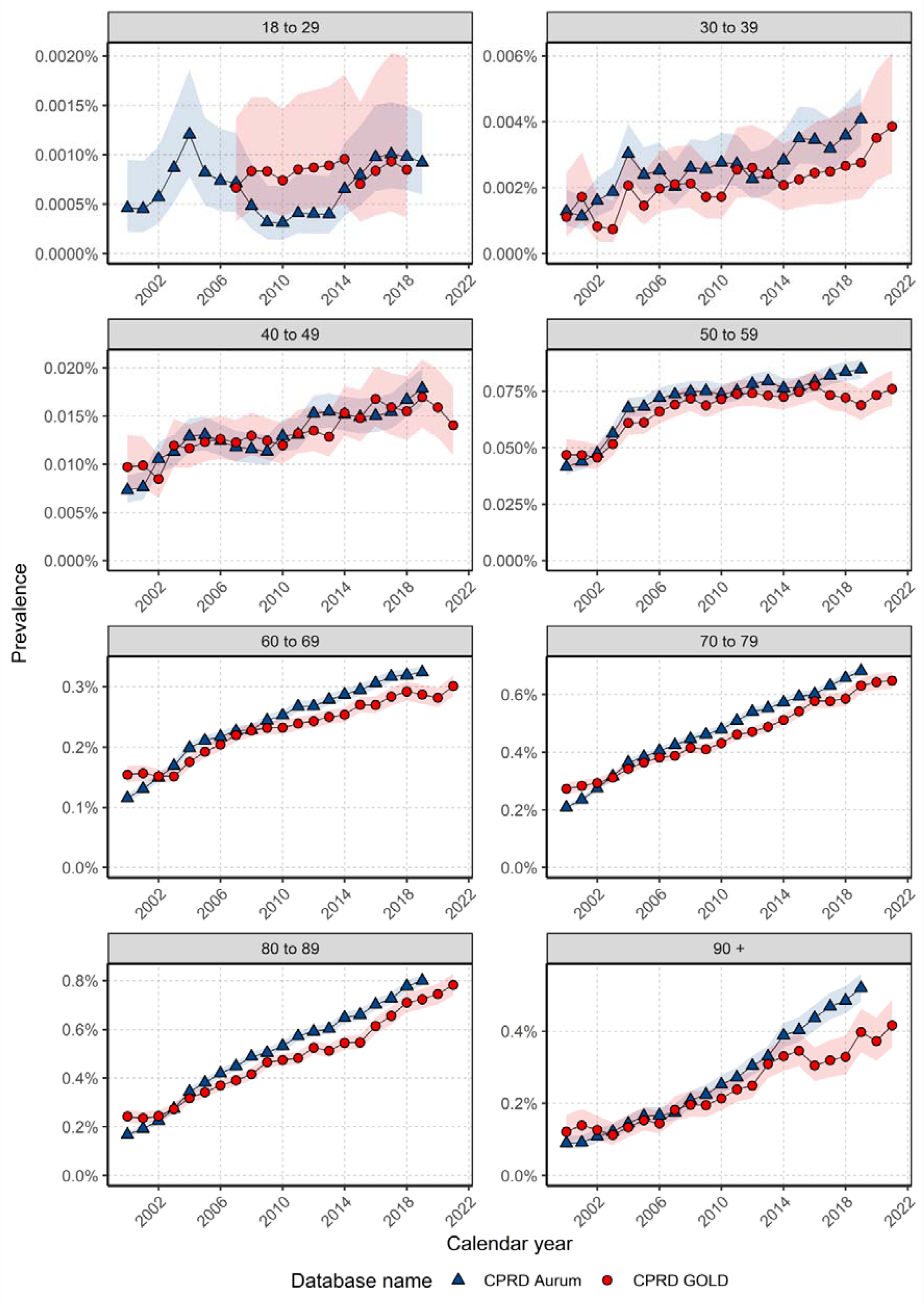
A**n**nual **prevalence from 2000 to 2021 stratified by database and age group. Bands show 95% confidence interval**.

Stratification by sex and age showed similar trends between males and females across age groups (Supplement S6). For those aged 50-59 years of age, PP for males remained relatively stable over the study period from 2005, whereas PP for females increased. For those 60-69 years of age, PP increased for females from 2010, whereas for males in this age group PP initially increased from 2010 to then decrease from 2014 in GOLD (in Aurum PP continued to increase, albeit at a slower rate than in females). Overall, males had higher PP compared to females for those aged 80 years and older. For those aged 50-79 years males had higher PP earlier in the study period with females having higher PP towards the end of the study period. For those aged 40-59 years, there were no differences in PP over time between males and females.

### 3.4 Overall survival rates for the cancer population with age, sex, and calendar year stratification

For GOLD there were 43,903 patients with 35,381 deaths (80.6% of patients) over the study period with a median follow-up of 0.54 years (IQR 0.18 – 1.39). For Aurum, there were 86,710 patients with 67,421 deaths (77.8% of patients) over the study period with a median follow-up of 0.58 years (IQR 0.19 – 1.49).

Supplement S7 shows survival curves for the overall population and stratified by sex. The median survival for the whole population was 0.66 years (95% CI 0.64 – 0.67) and 0.71 years (95% CI 0.70 – 0.72) in GOLD and Aurum respectively. Survival after one, five and ten-years after diagnosis was 39.0%, 12.0% and 6.4% for GOLD with similar values for Aurum. Median survival was higher in females (0.75-0.81 years) compared to males (0.60-0.64 years) across both databases. Regarding short– and long-term survival, females had better survival compared with males (Supplement S8).

When stratifying by age group, median survival decreased with age for both databases from 1.3-1.5 years for those aged 30-39 years to ∼0.35 years for those aged 90 years and older (Supplement S9). Survival at one, five, and ten-years overall decreased with increasing age with similar results across both databases (Supplement S10).

Median survival increased from 6.6 months in those diagnosed between 2000 to 2004 to 10 months for those diagnosed between 2015 to 2019 with a similar observation found for males and females (Figure 6, Supplement S11). Median survival was similar to those diagnosed in 2020-2021 to 2015-2019. For different age groups, median survival increased from those diagnosed in 2000-2004 to 2015-2019 in most age groups (40-89 years of age) apart from those 90 years and older where median survival has not changed over time.

**Figure 6:**
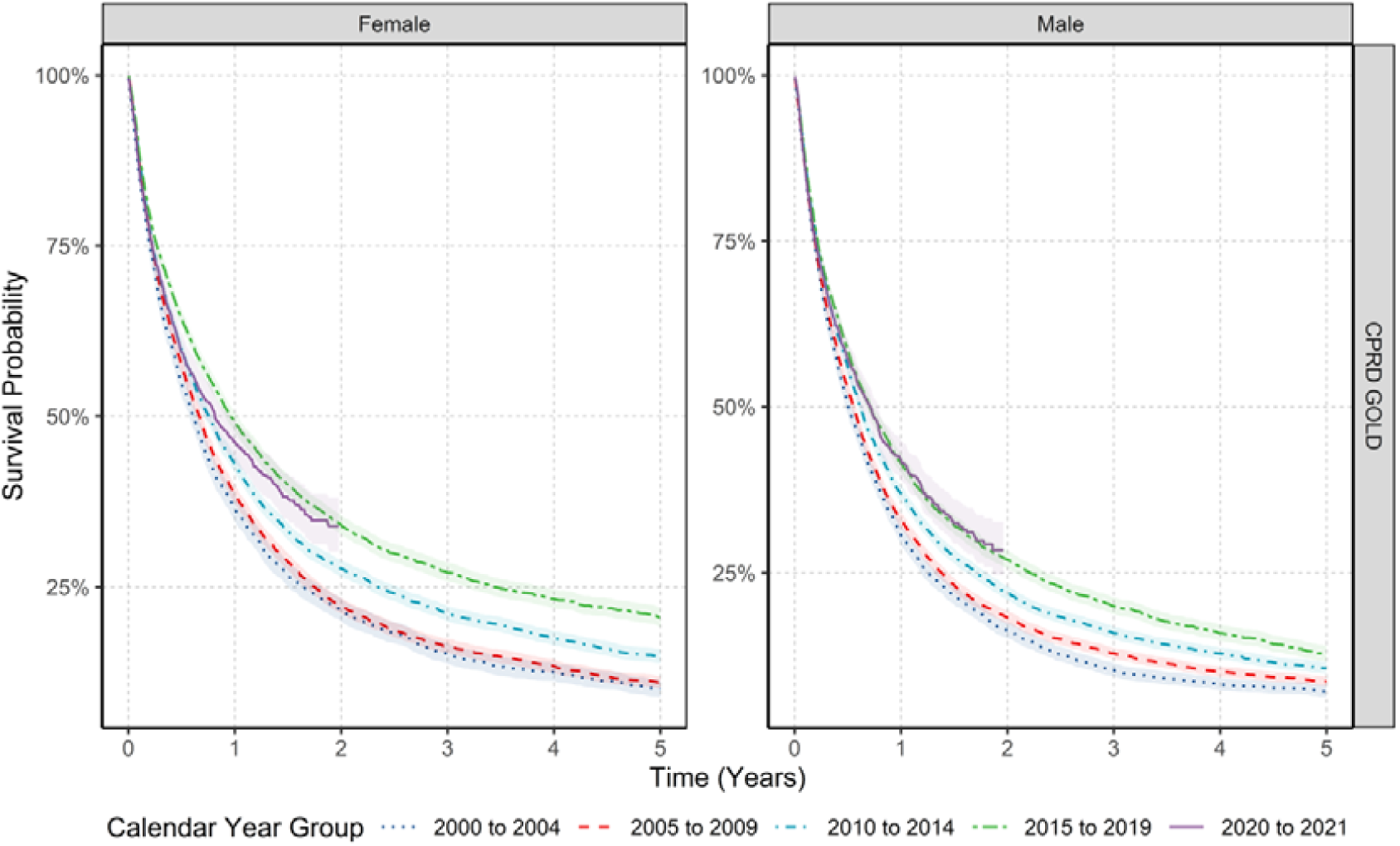
K**a**plan**-Meier survival curve of lung cancer stratified by sex and calendar year of diagnosis (2000-2004, 2005-2009, 2010-2014, 2015-2019 and 2020-2021). Bands show 95% confidence interval.**

Overall, one-year survival has increased between 2000-2004 and 2015-2019 for the whole population with similar trends for both sexes for GOLD. For the whole population, one-year survival in 2000-2004 increased from 33% to 45% in 2015-2019 with linear increases from 2005-2009 to 2015-2019. One-year survival in those diagnosed in 2020-2021 was similar to one-year survival for those diagnosed in 2015-2019 in GOLD. For five-year survival, there were linear increases in survival over time for GOLD from 2000-2009 to 2015-2019 (Supplement S12). When stratifying by age group, both one– and five-year survival were higher in those diagnosed between 2015-2019 compared to those diagnosed between 2000-2004 for those 50-89 years of age with similar patterns for both sexes.

## 4. DISCUSSION

This study provides a comprehensive analysis of trends in lung cancer incidence, prevalence, and survival in the UK. In this large cohort of over 11 million people, the incidence and prevalence of lung cancers in the UK has increased from 2000 to 2021, while both short-term and long-term survival has slightly improved across all age groups over this time period, with better survival in females than males.

Incidence rates reported here are broadly in line with National Cancer Statistics and Rapid Cancer Registration Databases[23], [24], [25]. Furthermore, for age and sex, studies have reported higher incidence rates in males compared to females and higher incidence with increasing age peaking in those aged 80 to 89 years of age in line with our estimates. Other studies have also shown increases in prevalence, particularly in females, where prevalence has overtaken males in recent years[1], [26], [27].

The increase in incidence and prevalence of lung cancer over time has been reported particularly in countries with higher levels of economic development as well as countries with higher smoking prevalence and air pollution[1], [2], [28]. These increases have largely been driven by females. However, other studies show a decline or stabilisation of the incidence of lung cancer due to decreases in males[1], [29], [30]. Incidence rates are known to vary by age as shown by this work and this is concordant with national cancer statistics[24] which show decreases in incidence over time in those aged 50-59 and aged over 80 particularly after 2014 with increases in those aged 60-79 years. These age differences in time trends are likely due to birth cohort effects as well as differences in modifiable risk factor exposures[24]. However, other studies also show incidence rates have decreased over time in all age groups which could be driven by a larger decrease in male incidence over time even with the increasing incidence in females[2], [31].

Encouragingly, despite the increases in incidence and prevalence, our study shows one– and five-year survival has improved over the past 20 years, with increases of 11% and 7% respectively. Our results are in line with the 2023 National Lung Cancer Audit for England and Wales[32], UK cancer registry[33], as well as other international studies[1], [34], [35], [36]. However, despite these improvements, short– and long-term survival is still low compared with other common cancers such as breast and prostate. Interestingly, five-year survival rates in this study are broadly comparable across Europe with values between 10-20%[1], [34], [35], [36].

The rise in lung cancer could be for numerous reasons, making future trends difficult to predict. Smoking is the greatest risk factor for the disease, with a 20-year delay between smoke exposure and cancer onset[37]. In our study, a high proportion of lung cancer patients (74%) were either current or ex-smokers; which is approximately five times higher than the overall population smoking prevalence[8] but slightly lower than UK national audit data estimates of smoking status in people with lung cancer[32]. However, following numerous successful public health interventions, the prevalence of smoking has fallen in the UK[8] as well as many other countries across Europe[38], although may have increased during recent lockdowns[39]. Furthermore, UK legislation has reduced the impact of ‘second-hand’ smoking which has amplified the impact of the reduction in smoking prevalence further in the UK[40]. Therefore, increases in disease burden in this study cannot be attributed to smoking prevalence alone. However, in our study, we further see an overall shift, from 2001-2021, in the age distribution of incident diagnoses, from younger to older participants – with incidence rising especially in females aged over 50, and males over 80. This may reflect that lower exposure to cigarette smoke is causing people that are predisposed to developing lung cancer, to develop it at an older age.

Alternatively, in recent years, electronic cigarettes have been promoted as a harm reduction strategy to a greater extent than in many other countries. The effects of this are largely unknown, especially with the lag time before cancer onset, so the effect of this may not yet be full seen[41].

Other risk factors include chronic obstructive pulmonary disease (COPD), which is an independent risk factor for lung cancer[2]. Nearly a quarter (24.5%) of patients had a prior diagnosis of COPD across both sexes, which is higher than existing estimates of COPD prevalence in the overall population[42].

In never-smokers, defined as individuals who have smoked fewer than 100 cigarettes in their lifetime[38], radon is the leading environmental cause of lung cancer[43]. As with smoking, there is a substantial exposure-cancer-mortality delay[44], and public health campaigns have aimed to reduce exposure[6]. Air pollution is also an established risk factor for lung cancer[45] with a study using UK Biobank demonstrating additive interactions between air pollution and genetic-related risk factors for lung cancer[46]. Despite apparent reductions of exposure to the discussed major modifiable risk factors in the UK, incidence and prevalence of lung cancer has increased in our study.

While not a sex-specific cancer, lung cancer shows sex-specific trends in our data as well as has been reported nationally and internationally[47][48], [49]. A recent review attributed this finding to many factors such as the slower relative reduction in female smoking compared to males[8], [48], coupled with females being exposed to different risk factors, as well as differences in oestrogen levels[48], exposure to human papillomavirus (HPV)[50] and genetic polymorphisms[48]. Furthermore, females have been reported to have a higher frequency of mutations in critical genes, such as TP53 and the KRAS leading to higher risk of the disease[51], [52], [53].

Although incidence and prevalence are increasing in females, overall survival is better in females compared to males in line with a similar study using primary care data from the UK[27]. Again, reasons for this difference are likely to be multifactorial involving different risk factors, treatment decisions, and cancer histology[47]. Survival from lung cancer has substantially improved since 2000, which has corresponded with major advancements in lung cancer treatments[2], [12]. Towards the start of this period, management of this disease progressively included a multi-disciplinary team (MDT) for cancer care[11], [54], [55]. Additionally, lung cancer treatments have further shifted towards more targeted regimens, after demonstrating better survival than existing therapy in trials [12]. Additionally, there has been a concurrent expansion of lung cancer surgery, which has been shown to further improve treatment outcomes [56], [57].

The decrease in survival but increase in incidence and prevalence in this study could also be due to better diagnostic methods and improved public awareness of lung cancer symptoms, leading to earlier detection. For instance, a 2012 UK Department of Health campaign was implemented to raise awareness of persistent cough as a lung cancer symptom, leading to a 3.1% increase in the proportion of NSCLC diagnosed at stage one[58], with similar campaigns since[59]. If diagnoses are indeed occurring at an earlier stage, lead-time bias may result in improvements in the survival data that do not exist in practice due to the fact the cancer was simply detected earlier, even if the treatment given and ultimate date of death is unchanged[60]. Of course, it may be the case that whilst cancers are diagnosed sooner, this is coupled with better treatment, not only for an equivalent stage due to improved therapy but also because even better treatment may be delivered at the earlier stage of cancer.

The main strength of this study is the use of a large primary care database covering the whole of the UK and validation of the results using another database from England. CPRD GOLD covers primary care practices from England, Wales, Scotland, and Northern Ireland whereas CPRD Aurum covers primary care practices in England. The similarity between the results in both databases, and their overall agreement with NCS and national audit programmes provide increased generalizability across the UK. The sharp increase in incidence at the start of the study period between 2000-2004 could be due and the institution of cancer quality improvement measures by the National Health Service in 2003 as well as the introduction of the Quality and Outcomes Framework (QOF) in 2004 which encourages general practitioners to record all new cases of cancer which could partly explain the increase in cancer recording[61].

Another strength of our study is the inclusion of a complete study population database for the assessment of incidence and prevalence. In contrast, cancer registry studies extrapolate the registry data to the whole population using national population statistics, potentially introducing biases[62], [63]. The high validity and completeness of mortality data with over 98% accuracy compared to national mortality records[20] allowed us to examine the impact of calendar time on overall survival – one of the key outcomes in cancer care.

Our study also has some limitations. Firstly, we used primary care data without linkage to a cancer registry which could lead to misclassification and delayed recording of cancer diagnoses[64]. However, previous validation studies have shown high accuracy and completeness of cancer diagnoses in primary care records[65]. Secondly, our use of primary care records also precluded us from studying tumour histology, genetic mutations, staging or cancer therapies, which can all impact lung cancer survival. Therefore, our survival estimates may overestimate survival in those with higher staging as well as those with specific subtypes or mutations associated with poorer survival such as SCLC[66]. Other factors, such as socio-economic status, environmental exposures such as air pollution and ethnicity could also result in different values for incidence, prevalence, and survival[45], [46]. Thirdly, in this study, we calculated overall survival, which does not differentiate between deaths caused by cancer vs. other causes. Therefore, it is a broad measure of overall survival rather than specifically cancer mortality. However, with the introduction of low-dose CT screening being introduced in the UK for targeted groups, the use of overall survival will enable the assessment of how much screening promotes overall survival and prevents all-cause mortality (including for instance changes related to non-lung cancer incidental diagnoses on CT screening), not just mortality related to lung cancer, and the overall benefit (or harm) associated with lung cancer treatments. Finally, smoking status was missing in 34.8% of lung cancer patients in this study with 48.3% of patients recorded as smokers which is around 3-5x the overall population prevalence of smoking for this period, however may be higher[8]

## 5. CONCLUSION

Despite the falling prevalence of smoking in the UK, the incidence and prevalence of lung cancer is increasing. Reassuringly, the improvements in survival over the study period highlight the development of better-targeted treatments and earlier diagnosis of high-risk populations. However, the rise of lung cancer even with a decrease in smoking is a cause for concern, particularly in females. Further work needs to focus on understanding this demographic shift, to explain why lung cancer continues to rise, which could lead to better prevention, earlier diagnosis, and further targeted treatments. As the UK Lung Cancer Screening Programme is introduced, with a target of 100% implementation by 2030, our study has the potential to enable subsequent comparisons of not only survival rates, but the baseline medical characteristics of people at diagnosis ultimately enabling a more comprehensive assessment of the impact of the screening programme and resulting public health interventions in the UK.

## Data Availability

All data produced in the present work are contained in the manuscript, included url links, and supplementary data.

## ABBREVIATIONS

IR: incidence rate
PP: period prevalence

## ACKNOWLEDMENTS CONTRIBUTIONS

All authors were involved in the study conception and design, interpretation of the results, and the preparation of the manuscript. DN carried out data analysis for the manuscript. AG, IT, developed and/or reviewed the clinical code lists used in this study and provided clinical expertise in this study. GC and DN wrote the initial draft of the manuscript with DPA. DN, EB, and DPA had access to the CPRD data. AD and WYM mapped the CPRD data to the OMOP CDM. All authors critically reviewed the final manuscript and gave consent for publication.

## FUNDING

This activity under the European Health Data & Evidence Network (EHDEN) and OPTIMA has received funding from the Innovative Medicines Initiative 2 (IMI2) Joint Undertaking under grant agreement No 806968 and No. 101034347 respectively. IMI2 receives support from the European Union’s Horizon 2020 research and innovation programme and European Federation of Pharmaceutical Industries and Associations (EFPIA). The sponsors of the study did not have any involvement in the writing of the manuscript or the decision to submit it for publication. Additionally, there was partial support from the Oxford NIHR Biomedical Research Centre. The corresponding author had full access to all the data in the study and had final responsibility for the decision to submit for publication.

## CONFLICTS OF INTEREST

Professor Daniel Prieto-Alhambra research group has received research grants from the European Medicines Agency, from the Innovative Medicines Initiative, from Amgen, Chiesi, and from UCB Biopharma; and consultancy or speaker fees (paid to his department) from Astellas, Amgen, Astra Zeneca, and UCB Biopharma. NB receives consultancy fees from Theramex and Sleep Universal Limited. All other authors declare no conflicts of interest.

## DATA AVAILIABILITY

This study is based in part on data from the Clinical Practice Research Datalink (CPRD) obtained under licence from the UK Medicines and Healthcare products Regulatory Agency. The data is provided by patients and collected by the NHS as part of their care and support. The interpretation and conclusions contained in this study are those of the author/s alone. Patient level data used in this study was obtained through an approved application to the CPRD (application number 22_001843) and is only available following an approval process to safeguard the confidentiality of patient data. Details on how to apply for data access can be found at https://cprd.com/data-access.

## Footnotes

1 CPRD: Clinical Practice Research Datalink. IR: Incidence rate. PP: Period prevalence.

## Notes

### Author Declarations

The protocol for this research was approved by the Research Data Governance (RDG) Board of the Medicine and Healthcare products Regulatory Agency database research (protocol number 22_001843).

