## Supplementary Data for "Incidence, prevalence, and survival of lung cancer in the United Kingdom from 2000-2021: a population-based cohort study"

### **S1: Clinical codelists for lung cancer**

The clinical codelists used for lung cancer is listed in the table below with the corresponding SNOMED concept ID, OMOP concept ID and concept description. Only diagnosis records alone were used to identify cancer outcome for this study. Different codelists were created for incident and prevalent definitions of lung cancer. We developed concept definitions using ATLAS, the OHDSI open-source platform (<https://github.com/OHDSI/atlas>). Clinical adjudicators reviewed the cohort definitions and associated concept sets.

| **Concept Id** | **Concept SNOMED Code** | **Concept Description** | **Code used for** |
| --- | --- | --- | --- |
| 443388 | 363358000 | Malignant tumor of lung | Incidence and prevalence |
| 258369 | 93880001 | Primary malignant neoplasm of lung | Incidence and prevalence |
| 4092216 | 187862007 | Malignant neoplasm of upper lobe of lung | Incidence and prevalence |
| 4089756 | 187870002 | Malignant neoplasm of lower lobe of lung | Incidence and prevalence |
| 4151250 | 269464000 | Malignant neoplasm of upper lobe, bronchus or lung | Incidence and prevalence |
| 4246121 | 93827000 | Primary malignant neoplasm of hilus of lung | Incidence and prevalence |
| 4089754 | 187866005 | Malignant neoplasm of middle lobe of lung | Incidence and prevalence |
| 4092217 | 187864008 | Malignant neoplasm of middle lobe, bronchus or lung | Incidence and prevalence |
| 258375 | 109371002 | Overlapping malignant neoplasm of bronchus and lung | Incidence and prevalence |
| 4092218 | 187865009 | Malignant neoplasm of middle lobe bronchus | Incidence and prevalence |
| 256646 | 372112000 | Primary malignant neoplasm of middle lobe, bronchus or lung | Incidence and prevalence |
| 261236 | 372135002 | Primary malignant neoplasm of upper lobe, bronchus or lung | Incidence and prevalence |
| 433973 | 93729006 | Primary malignant neoplasm of bronchus of left lower lobe | Incidence and prevalence |
| 762426 | 354701000119107 | Primary malignant neoplasm of left lung | Incidence and prevalence |
| 762427 | 354741000119109 | Primary malignant neoplasm of right lung | Incidence and prevalence |
| 4110587 | 254625005 | Malignant tumor of lung parenchyma | Incidence and prevalence |
| 4110589 | 254629004 | Large cell carcinoma of lung | Incidence and prevalence |
| 4110590 | 254631008 | Giant cell carcinoma of lung | Incidence and prevalence |
| 4110591 | 254632001 | Small cell carcinoma of lung | Incidence and prevalence |
| 4110705 | 254634000 | Squamous cell carcinoma of lung | Incidence and prevalence |
| 4110706 | 254638002 | Pancoast tumor | Incidence and prevalence |
| 4111807 | 254635004 | Epithelioid hemangioendothelioma of lung | Incidence and prevalence |
| 4112738 | 254626006 | Adenocarcinoma of lung | Incidence and prevalence |
| 4112739 | 254633006 | Oat cell carcinoma of lung | Incidence and prevalence |
| 4115276 | 254637007 | Non-small cell lung cancer | Incidence and prevalence |
| 4140471 | 427038005 | Epidermal growth factor receptor negative non-small cell lung cancer | Incidence and prevalence |
| 4143825 | 426964009 | Epidermal growth factor receptor positive non-small cell lung cancer | Incidence and prevalence |
| 4155293 | 372111007 | Carcinoma of lower lobe, bronchus or lung | Incidence and prevalence |
| 4157454 | 372110008 | Primary malignant neoplasm of lower lobe, bronchus or lung | Incidence and prevalence |
| 4162248 | 372113005 | Carcinoma of middle lobe, bronchus or lung | Incidence and prevalence |
| 4162252 | 372136001 | Carcinoma of upper lobe, bronchus or lung | Incidence and prevalence |
| 4196724 | 313353007 | Squamous cell carcinoma of bronchus in left lower lobe | Incidence and prevalence |
| 4196725 | 313355000 | Squamous cell carcinoma of bronchus in right lower lobe | Incidence and prevalence |
| 4197581 | 313354001 | Squamous cell carcinoma of bronchus in left upper lobe | Incidence and prevalence |
| 4197582 | 313356004 | Squamous cell carcinoma of bronchus in right middle lobe | Incidence and prevalence |
| 4197583 | 313357008 | Squamous cell carcinoma of bronchus in right upper lobe | Incidence and prevalence |
| 4208307 | 440173001 | Nonsquamous nonsmall cell neoplasm of lung | Incidence and prevalence |
| 4246027 | 93730001 | Primary malignant neoplasm of bronchus of left upper lobe | Incidence and prevalence |
| 4246126 | 93865007 | Primary malignant neoplasm of left upper lobe of lung | Incidence and prevalence |
| 4246148 | 93991009 | Primary malignant neoplasm of right lower lobe of lung | Incidence and prevalence |
| 4246804 | 93732009 | Primary malignant neoplasm of bronchus of right middle lobe | Incidence and prevalence |
| 4246805 | 93733004 | Primary malignant neoplasm of bronchus of right upper lobe | Incidence and prevalence |
| 4247727 | 93731002 | Primary malignant neoplasm of bronchus of right lower lobe | Incidence and prevalence |
| 4247832 | 93864006 | Primary malignant neoplasm of lower lobe of left lung | Incidence and prevalence |
| 4307118 | 423050000 | Large cell carcinoma of lung, TNM stage 2 | Incidence and prevalence |
| 4308479 | 423121009 | Non-small cell carcinoma of lung, TNM stage 4 | Incidence and prevalence |
| 4310448 | 423295000 | Squamous cell carcinoma of lung, TNM stage 1 | Incidence and prevalence |
| 4310703 | 424132000 | Non-small cell carcinoma of lung, TNM stage 1 | Incidence and prevalence |
| 4311501 | 93992002 | Primary malignant neoplasm of right middle lobe of lung | Incidence and prevalence |
| 4311997 | 422968005 | Non-small cell carcinoma of lung, TNM stage 3 | Incidence and prevalence |
| 4312274 | 425376008 | Squamous cell carcinoma of lung, TNM stage 4 | Incidence and prevalence |
| 4312567 | 93993007 | Primary malignant neoplasm of upper lobe of right lung | Incidence and prevalence |
| 4312768 | 424970000 | Large cell carcinoma of lung, TNM stage 3 | Incidence and prevalence |
| 4313200 | 423468007 | Squamous cell carcinoma of lung, TNM stage 2 | Incidence and prevalence |
| 4313751 | 423600008 | Large cell carcinoma of lung, TNM stage 4 | Incidence and prevalence |
| 4314040 | 424938000 | Large cell carcinoma of lung, TNM stage 1 | Incidence and prevalence |
| 4314172 | 425048006 | Non-small cell carcinoma of lung, TNM stage 2 | Incidence and prevalence |
| 4322387 | 425230006 | Squamous cell carcinoma of lung, TNM stage 3 | Incidence and prevalence |
| 36686537 | 12235561000119100 | Large cell carcinoma of left lung | Incidence and prevalence |
| 36686538 | 12235601000119100 | Large cell carcinoma of right lung | Incidence and prevalence |
| 36712707 | 1078881000119100 | Primary adenocarcinoma of lower lobe of left lung | Incidence and prevalence |
| 36712708 | 1078901000119100 | Primary adenocarcinoma of upper lobe of left lung | Incidence and prevalence |
| 36712709 | 1078931000119100 | Primary adenocarcinoma of lower lobe of right lung | Incidence and prevalence |
| 36712815 | 12240951000119100 | Squamous cell carcinoma of left lung | Incidence and prevalence |
| 36712816 | 12240991000119100 | Squamous cell carcinoma of right lung | Incidence and prevalence |
| 36712981 | 15956381000119100 | Adenocarcinoma of right lung | Incidence and prevalence |
| 36713366 | 683991000119103 | Extensive stage primary small cell carcinoma of lung | Incidence and prevalence |
| 36716426 | 722425009 | Reactive oxygen species 1 positive non-small cell lung cancer | Incidence and prevalence |
| 36716500 | 722528008 | Primary malignant neuroendocrine neoplasm of lung | Incidence and prevalence |
| 36717017 | 1078961000119100 | Primary adenocarcinoma of upper lobe of right lung | Incidence and prevalence |
| 37109576 | 723301009 | Squamous non-small cell lung cancer | Incidence and prevalence |
| 37110031 | 724056005 | Malignant neoplasm of lower lobe of right lung | Incidence and prevalence |
| 37110032 | 724058006 | Malignant neoplasm of upper lobe of left lung | Incidence and prevalence |
| 37110033 | 724059003 | Malignant neoplasm of lower lobe of left lung | Incidence and prevalence |
| 37110034 | 724060008 | Malignant neoplasm of right upper lobe of lung | Incidence and prevalence |
| 37311684 | 822969007 | Acinar cell cystadenocarcinoma of lung | Incidence and prevalence |
| 37395648 | 67811000119102 | Primary small cell malignant neoplasm of lung, TNM stage 1 | Incidence and prevalence |
| 37395649 | 67821000119109 | Primary small cell malignant neoplasm of lung, TNM stage 2 | Incidence and prevalence |
| 37395650 | 67831000119107 | Primary small cell malignant neoplasm of lung, TNM stage 3 | Incidence and prevalence |
| 37395651 | 67841000119103 | Primary small cell malignant neoplasm of lung, TNM stage 4 | Incidence and prevalence |
| 40391740 | 189815007 | Pulmonary blastoma | Incidence and prevalence |
| 40492938 | 448993007 | Carcinoma of lung | Incidence and prevalence |
| 42539251 | 15956341000119100 | Adenocarcinoma of left lung | Incidence and prevalence |
| 45766129 | 703228009 | Non-small cell lung cancer with mutation in epidermal growth factor receptor | Incidence and prevalence |
| 45766131 | 703230006 | Non-small cell lung cancer without mutation in epidermal growth factor receptor | Incidence and prevalence |
| 45768879 | 707403002 | Primary fetal adenocarcinoma of lung | Incidence and prevalence |
| 45768880 | 707404008 | Primary mixed subtype adenocarcinoma of lung | Incidence and prevalence |
| 45768881 | 707405009 | Primary adenosquamous carcinoma of lung | Incidence and prevalence |
| 45768883 | 707408006 | Primary small cell non-keratinizing squamous cell carcinoma of lung | Incidence and prevalence |
| 45768884 | 707409003 | Primary acinar cell carcinoma of lung | Incidence and prevalence |
| 45768885 | 707410008 | Primary solid carcinoma of lung | Incidence and prevalence |
| 45768886 | 707411007 | Primary papillary adenocarcinoma of lung | Incidence and prevalence |
| 45768916 | 707451005 | Primary adenocarcinoma of lung | Incidence and prevalence |
| 45768917 | 707452003 | Primary mucinous adenocarcinoma of lung | Incidence and prevalence |
| 45768918 | 707453008 | Primary clear cell squamous cell carcinoma of lung | Incidence and prevalence |
| 45768919 | 707454002 | Primary basaloid squamous cell carcinoma of lung | Incidence and prevalence |
| 45768920 | 707456000 | Primary undifferentiated carcinoma of lung | Incidence and prevalence |
| 45768921 | 707457009 | Primary spindle cell carcinoma of lung | Incidence and prevalence |
| 45768922 | 707458004 | Primary pleomorphic carcinoma of lung | Incidence and prevalence |
| 45768923 | 707460002 | Primary pseudosarcomatous carcinoma of lung | Incidence and prevalence |
| 45768927 | 707464006 | Primary myoepithelial carcinoma of lung | Incidence and prevalence |
| 45768928 | 707466008 | Primary adenoid cystic carcinoma of lung | Incidence and prevalence |
| 45768929 | 707467004 | Primary salivary gland type carcinoma of lung | Incidence and prevalence |
| 45768930 | 707468009 | Primary mixed mucinous and non-mucinous bronchiolo-alveolar carcinoma of lung | Incidence and prevalence |
| 45768931 | 707469001 | Primary non-mucinous bronchiolo-alveolar carcinoma of lung | Incidence and prevalence |
| 45768932 | 707470000 | Primary mucinous bronchiolo-alveolar carcinoma of lung | Incidence and prevalence |
| 45769034 | 707595001 | Primary mucinous cystadenocarcinoma of lung | Incidence and prevalence |
| 45769035 | 707596000 | Primary carcinosarcoma of lung | Incidence and prevalence |
| 45772933 | 707407001 | Primary signet ring cell carcinoma of lung | Incidence and prevalence |
| 45772938 | 707455001 | Primary papillary squamous cell carcinoma of lung | Incidence and prevalence |
| 45772939 | 707465007 | Primary mucoepidermoid carcinoma of lung | Incidence and prevalence |
| 46272955 | 711414003 | Primary clear cell adenocarcinoma of lung | Incidence and prevalence |
| 4198434 | 314954002 | Local recurrence of malignant tumor of lung | Prevalence only |
| 4201621 | 315006004 | Metastasis from malignant tumor of lung | Prevalence only |

### **S2: Population attrition showing eligible patients for study from each database**

| **N** | **Reason** | **N excluded** | **Database** |
| --- | --- | --- | --- |
| 39999011 | Starting population |  | Aurum |
| 39999011 | Missing year of birth | 0 |  |
| 39999011 | Missing sex | 0 |  |
| 34833388 | Cannot satisfy age criteria during the study period based on year of birth | 5165623 |  |
| 29190480 | No observation time available during study period | 5642908 |  |
| 29190480 | Doesn't satisfy age criteria during the study period | 0 |  |
| 25483313 | Prior history requirement not fulfilled during study period | 3707167 |  |
| 24340860 | No observation time available after applying age and prior history criteria | 1142453 |  |
| 24340860 | Starting analysis population |  |  |
| 24340860 | Estimating prevalence |  |  |
| 24334950 | Excluded due to prior event (do not pass outcome washout during study period) | 5910 |  |
| 24334950 | Estimating incidence |  |  |
| 88540 | With a cancer diagnosis | 24246410 |  |
| 86710 | Cancer diagnosis not on same date as death | 1830 |  |
| 86710 | Estimating survival |  |  |
| 17054819 | Starting population |  | GOLD |
| 17054819 | Missing year of birth | 0 |  |
| 17054819 | Missing sex | 0 |  |
| 15210165 | Cannot satisfy age criteria during the study period based on year of birth | 1844654 |  |
| 13978229 | No observation time available during study period | 1231936 |  |
| 13978229 | Doesn't satisfy age criteria during the study period | 0 |  |
| 12254874 | Prior history requirement not fulfilled during study period | 1723355 |  |
| 11388117 | No observation time available after applying age and prior history criteria | 866757 |  |
| 11388117 | Starting analysis population |  |  |
| 11388117 | Estimating prevalence |  |  |
| 11385621 | Excluded due to prior event (do not pass outcome washout during study period) | 2496 |  |
| 11385621 | Estimating incidence |  |  |
| 45563 | With a cancer diagnosis | 11340058 |  |
| 43903 | Cancer diagnosis not on same date as death | 1660 |  |
| 43903 | Estimating survival |  |  |

### **S3: Baseline characteristics of lung cancer patients at the time of diagnosis for CPRD Aurum.**

| **Database** | **CPRD Aurum** |
| --- | --- |
| **Number of patients** | 88540 |
| **Sex: Male (N[%])** | 48819 (55.1%) |
| **Age (Median [IQR])** | 73 (65 to 80) |
| **Age Groups N (%)** |  |
| 18-29 | 45 (0.1%) |
| 30-39 | 274 (0.3%) |
| 40-49 | 1880 (2.1%) |
| 50-59 | 8981 (10.1%) |
| 60-69 | 23278 (26.3%) |
| 70-79 | 31897 (36.0%) |
| 80-89 | 19488 (22.0%) |
| 90+ | 2697 (3.00%) |
| **Prior history, days** |  |
| median [IQR] | 6775 (3,487 to 10,987) |
| **General conditions (any time prior)** |  |
| Atrial fibrillation | 7056 (8.0%) |
| Cerebrovascular disease | 8298 (9.4%) |
| Chronic liver disease | 530 (0.6%) |
| Chronic obstructive lung disease | 23599 (26.7%) |
| Coronary arteriosclerosis | 1261 (1.4%) |
| Crohn’s disease | 346 (0.4%) |
| Dementia | 1656 (1.9%) |
| Depressive disorder | 12974 (14.7%) |
| Diabetes mellitus | 12822 (14.5%) |
| Gastroesophageal reflux disease | 3058 (3.5%) |
| Gastrointestinal haemorrhage | 6691 (7.6%) |
| Heart disease | 24166 (27.3%) |
| Heart failure | 4276 (4.8%) |
| Hepatitis C | 126 (0.1%) |
| HIV | 23 (0.00%) |
| Hyperlipidemia | 10449 (11.8%) |
| Hypertensive disorder | 35469 (40.1%) |
| Ischemic heart disease | 14929 (16.9%) |
| Lesion of liver | 1556 (1.8%) |
| Obesity | 2403 (2.7%) |
| Osteoarthritis | 25083 (28.3%) |
| Peripheral vascular disease | 4932 (5.6%) |
| Pneumonia | 6741 (7.6%) |
| Psoriasis | 3740 (4.2%) |
| Pulmonary embolism | 1923 (2.2%) |
| Renal impairment | 13373 (15.1%) |
| Rheumatoid arthritis | 2104 (2.4%) |
| Schizophrenia | 420 (0.5%) |
| Ulcerative colitis | 472 (0.5%) |
| Urinary tract infectious disease | 12018 (13.6%) |
| Venous thrombosis | 5252 (5.9%) |
| Visual system disorder | 36552 (41.3%) |

### **S4: Overall incidence rates (with 95% confidence intervals) for lung cancer from 2000 to 2021 for CPRD GOLD and 2000 to 2019 for Aurum stratified by database and age group**

| **Age Group (years)** | **n persons** | **pys** | **n events** | **Incidence (100,000 pys)** | **Database** |
| --- | --- | --- | --- | --- | --- |
| 18 to 29 | 9,238,441 | 31,941,436 | 45 | 0.14 (0.10 to 0.19) | CPRD Aurum |
| 30 to 39 | 8,292,622 | 32,680,100 | 274 | 0.84 (0.74 to 0.94) |  |
| 40 to 49 | 6,515,966 | 32,924,342 | 1,880 | 5.71 (5.45 to 5.97) |  |
| 50 to 59 | 5,438,224 | 28,668,826 | 8,981 | 31.33 (30.7 to 32.0) |  |
| 60 to 69 | 4,170,276 | 22,500,850 | 23,278 | 103.5 (102.1 to 104.8) |  |
| 70 to 79 | 3,119,104 | 16,252,188 | 31,897 | 196.3 (194.1 to 198.4) |  |
| 80 to 89 | 1,949,102 | 8,834,297 | 19,488 | 220.59 (217.5 to 223.7) |  |
| 90 + | 666,317 | 2,269,618 | 2,697 | 118.8 (114.4 to 123.4) |  |
| 18 to 29 | 3,871,131 | 16,018,568 | 24 | 0.2 (0.1 to 0.2) | CPRD GOLD |
| 30 to 39 | 3,682,403 | 15,107,431 | 129 | 0.9 (0.7 to 1.0) |  |
| 40 to 49 | 3,246,901 | 16,116,689 | 1,030 | 6.4 (6.0 to 6.8) |  |
| 50 to 59 | 2,885,599 | 14,721,668 | 4,660 | 31.7 (30.8 to 32.6) |  |
| 60 to 69 | 2,293,348 | 11,892,162 | 12,249 | 103.0 (101.2 to 104.8) |  |
| 70 to 79 | 1,682,118 | 8,407,711 | 16,745 | 199.2 (196.2 to 202.2) |  |
| 80 to 89 | 1,023,695 | 4,422,107 | 9,546 | 215.9 (211.6 to 220.2) |  |
| 90 + | 322,801 | 965,457 | 1,181 | 122.3 (115.5 to 129.5) |  |

**Pys person years**

### **S5: Annualised incidence rates for lung cancer from 2000 to 2021 stratified by database, sex and age group. Band represents 95% CI.**

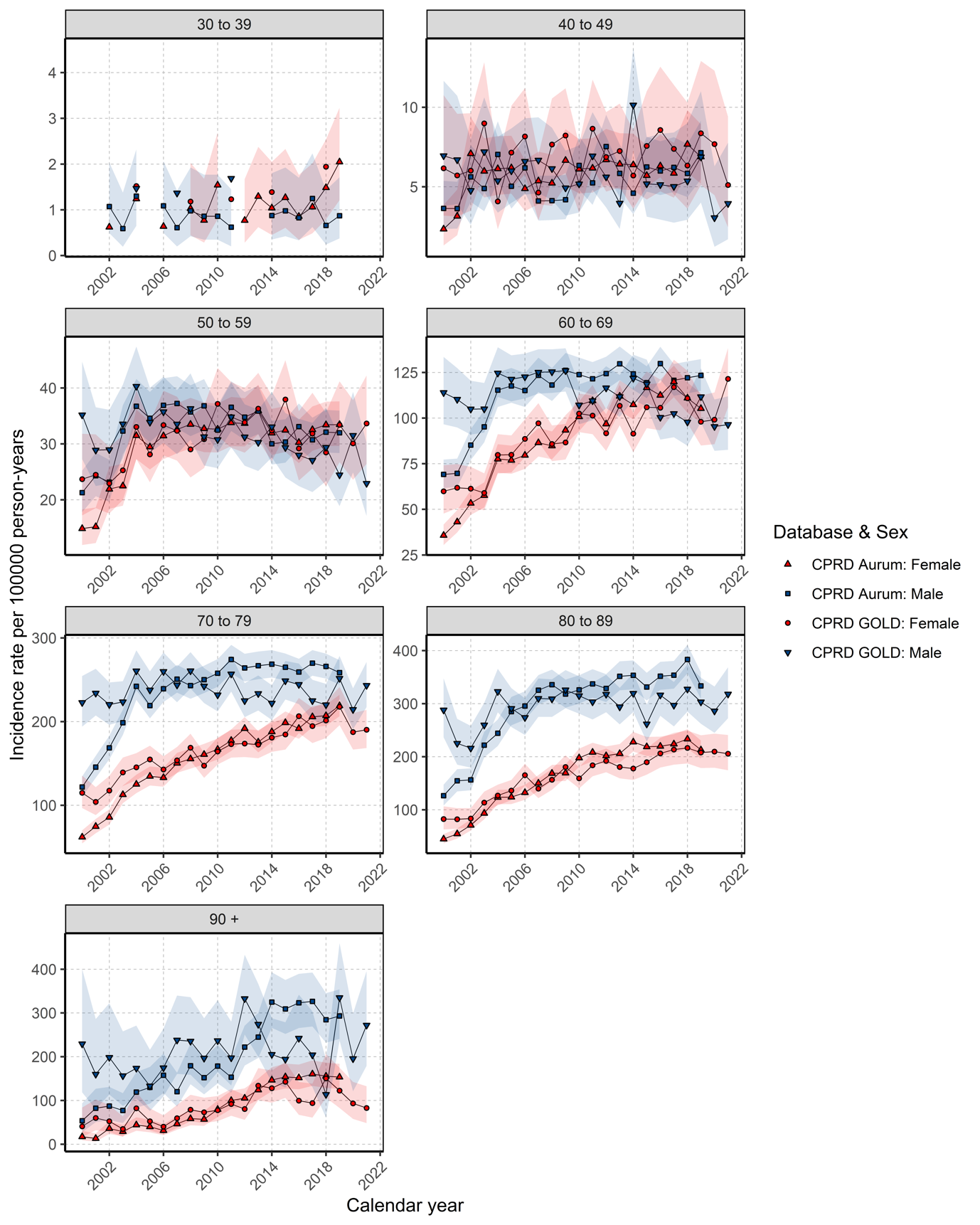

### **S6: Annualised prevalence for lung cancer from 2000 to 2021 stratified by database, sex and age group. Band represents 95% CI.**

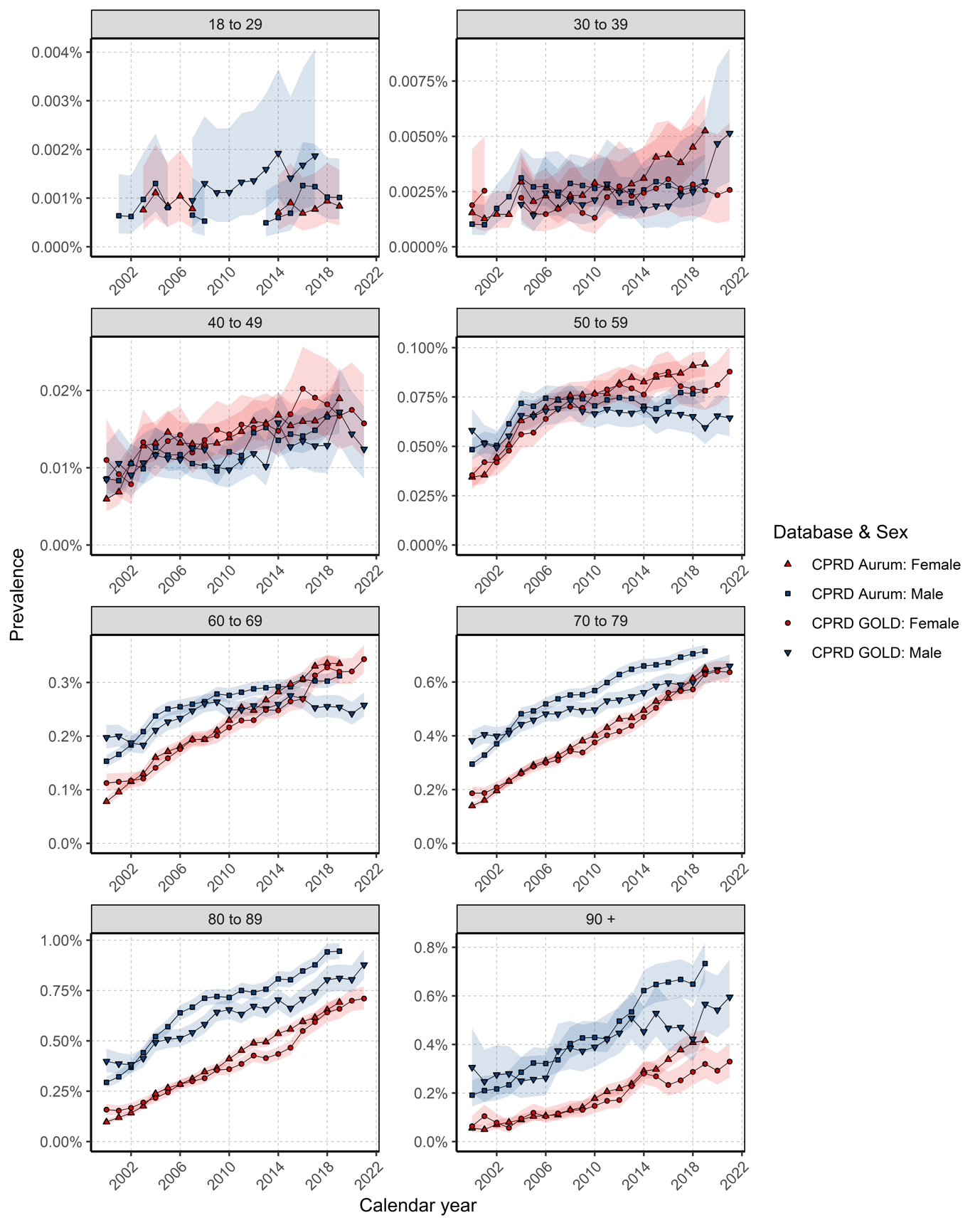

### **S7: Kaplan-Meier survival curve of lung cancer by database and sex.**

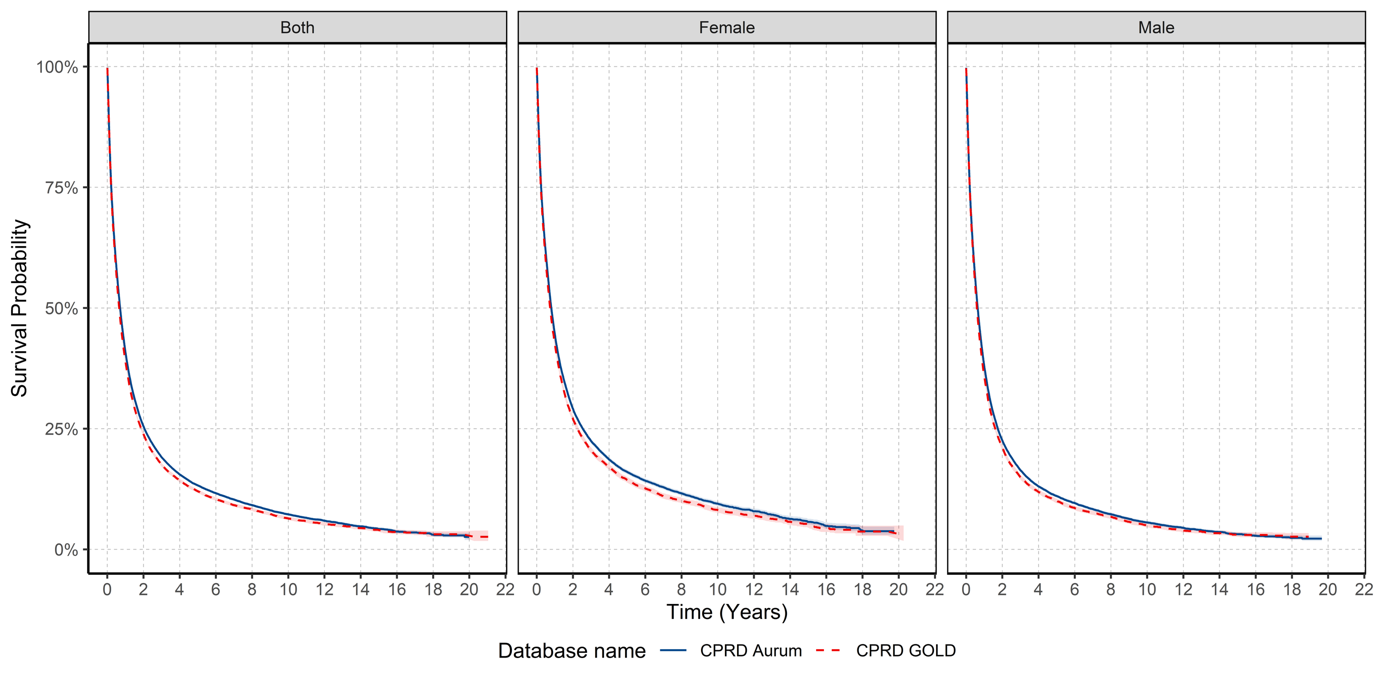

### **S8: Survival (%) after 1, 5 and 10 years after lung cancer diagnosis stratified by database and sex**

| **Time** | **Sex** | **% Survival (95% CI)** | **Database** |
| --- | --- | --- | --- |
| 1 | Male | 38.0 (37.6 - 38.5) | Aurum |
| 5 |  | 11.1 (10.7 - 11.4) |  |
| 10 |  | 5.6 (5.3 - 5.9) |  |
| 1 | Female | 44.6 (44.1 - 45.2) |  |
| 5 |  | 16.0 (15.6 - 16.5) |  |
| 10 |  | 9.46 (9.0 - 9.9) |  |
| 1 | Male | 35.8 (35.21 - 36.48) | GOLD |
| 5 |  | 10.0 (9.6 - 10.5) |  |
| 10 |  | 5.0 (4.6 - 5.4) |  |
| 1 | Female | 42.6 (41.9 - 43.4) |  |
| 5 |  | 14.4 (13.8 - 15.0) |  |
| 10 |  | 8.2 (7.6 - 8.78) |  |

CI: confidence interval

### **S9: Median survival stratified by database and age group**

| **Age Group (years)** | **Median survival in years (%95 CI)** | **n persons** | **n events** | **Database** |
| --- | --- | --- | --- | --- |
| 18 to 29 | Not achieved | 45 | 14 | Aurum |
| 30 to 39 | 1.314 (1.008 - 2.062) | 272 | 163 |  |
| 40 to 49 | 0.947 (0.862 - 1.038) | 1850 | 1323 |  |
| 50 to 59 | 0.917 (0.890 - 0.945) | 8845 | 6603 |  |
| 60 to 69 | 0.851 (0.830 - 0.871) | 22923 | 17474 |  |
| 70 to 79 | 0.698 (0.684 - 0.715) | 31247 | 24582 |  |
| 80 to 89 | 0.504 (0.485 - 0.523) | 18932 | 15166 |  |
| 90 + | 0.361 (0.329 - 0.397) | 2596 | 2096 |  |
| 18 to 29 | Not achieved | 24 | 6 | GOLD |
| 30 to 39 | 1.478 (0.810 - 2.177) | 127 | 82 |  |
| 40 to 49 | 0.903 (0.786 - 1.027) | 999 | 740 |  |
| 50 to 59 | 0.827 (0.789 - 0.873) | 4549 | 3467 |  |
| 60 to 69 | 0.797 (0.775 - 0.824) | 11907 | 9415 |  |
| 70 to 79 | 0.638 (0.621 - 0.657) | 16164 | 13232 |  |
| 80 to 89 | 0.463 (0.449 - 0.479) | 9041 | 7535 |  |
| 90 + | 0.353 (0.301 - 0.386) | 1092 | 904 |  |

Not achieved: Median survival was not achieved in study period.

### **S10: Survival rates (and 95% confidence intervals) of lung cancer from for GOLD (2000-2021) and Aurum (2000-2019) stratified by database and age group.**

| **Age Group (years)** | **One-year survival (%)** | | **Five-year survival (%)** | | **Ten-year survival (%)** | |
| --- | --- | --- | --- | --- | --- | --- |
|  | GOLD | Aurum | GOLD | Aurum | GOLD | Aurum |
| 18-29 | 82.7 (68.6 - 99.7) | 77.3 (65.3 - 91.7) | 71.6 (54.3 - 94.2) | 57.5 (40.4 - 81.6) | - | - |
| 30-39 | 54.0 (45.7 - 63.7) | 56.0 (50.1 - 62.6) | 30.3 (22.6 - 40.6) | 31.9 (26.1 - 39.1) | 21.6 (13.1 - 35.4) | 26.0 (20.1 - 33.5) |
| 40-49 | 47.8 (44.7 - 51.1) | 48.6 (46.3 - 51.1) | 19.4 (16.8 - 22.4) | 21.3 (19.3 - 23.6) | 14.8 (12.2 - 18.0) | 15.5 (13.4 - 17.9) |
| 50-59 | 44.7 (43.2 - 46.2) | 47.2 (46.1 - 48.3) | 16.4 (15.2 - 17.7) | 17.6 (16.7 - 18.5) | 11.3 (10.1 - 12.6) | 12.0 (11.1 - 13.0) |
| 60-69 | 43.5 (42.6 - 44.5) | 45.4 (44.7 - 46.0) | 15.1 (14.3 - 15.8) | 16.8 (16.2 - 17.3) | 8.6 (7.9 - 9.4) | 10.1 (9.49 - 10.7) |
| 70-79 | 38.4 (37.6 - 39.2) | 40.9 (40.3 - 41.4) | 11.3 (10.7 - 11.9) | 12.6 (12.1 - 13.0) | 5.1 (4.5 - 5.6) | 5.76 (5.34 - 6.22) |
| 80-89 | 31.5 (30.5 - 32.5) | 33.7 (33.0 - 34.4) | 6.2 (5.6 - 6.9) | 7.5 (7.0 - 8.0) | 1.5 (1.0 - 2.1) | 2.25 (1.83 - 2.75) |
| 90+ | 24.2 (21.6 - 27.0) | 25.6 (23.9 - 27.5) | 3.2 (1.9 - 5.5) | 2.53 (1.70 - 3.78) | - | 0.51 (0.17 - 1.50) |

### **S11: Median survival stratified by database, calendar year for whole population and sex**

| **Sex** | **Calendar Year** | **Median survival in years (95% CI)** | **n persons** | **n events** | **Database** |
| --- | --- | --- | --- | --- | --- |
| Both | 2000 to 2004 | 0.635 (0.613 - 0.654) | 12019 | 8028 | Aurum |
| Male |  | 0.591 (0.564 - 0.616) | 7188 | 4977 |  |
| Female |  | 0.701 (0.671 - 0.753) | 4831 | 3051 |  |
| Both | 2005 to 2009 | 0.594 (0.578 - 0.608) | 20204 | 14397 |  |
| Male |  | 0.561 (0.542 - 0.583) | 11592 | 8529 |  |
| Female |  | 0.646 (0.616 - 0.671) | 8612 | 5868 |  |
| Both | 2010 to 2014 | 0.690 (0.674 - 0.709) | 25364 | 16950 |  |
| Male |  | 0.624 (0.602 - 0.649) | 13678 | 9458 |  |
| Female |  | 0.772 (0.745 - 0.799) | 11686 | 7492 |  |
| Both | 2015 to 2019 | 0.923 (0.898 - 0.947) | 29123 | 17285 |  |
| Male |  | 0.783 (0.756 - 0.819) | 15311 | 9670 |  |
| Female |  | 1.125 (1.073 - 1.175) | 13812 | 7615 |  |
| Both | 2000 to 2004 | 0.559 (0.526 - 0.589) | 6250 | 4259 | GOLD |
| Male |  | 0.512 (0.482 - 0.548) | 3718 | 2617 |  |
| Female |  | 0.630 (0.586 - 0.679) | 2532 | 1642 |  |
| Both | 2005 to 2009 | 0.594 (0.578 - 0.613) | 12111 | 8617 |  |
| Male |  | 0.553 (0.534 - 0.578) | 6792 | 4963 |  |
| Female |  | 0.660 (0.627 - 0.687) | 5319 | 3654 |  |
| Both | 2010 to 2014 | 0.668 (0.643 - 0.693) | 12847 | 8616 |  |
| Male |  | 0.611 (0.591 - 0.643) | 6780 | 4691 |  |
| Female |  | 0.745 (0.706 - 0.786) | 6067 | 3925 |  |
| Both | 2015 to 2019 | 0.832 (0.791 - 0.865) | 9650 | 5951 |  |
| Male |  | 0.709 (0.665 - 0.745) | 4808 | 3099 |  |
| Female |  | 0.950 (0.898 - 1.021) | 4842 | 2852 |  |
| Both | 2020 to 2021 | 0.767 (0.690 - 0.810) | 3045 | 1490 |  |
| Male |  | 0.712 (0.619 - 0.789) | 1523 | 776 |  |
| Female |  | 0.816 (0.709 - 0.958) | 1522 | 714 |  |

CI: confidence interval

### **S12: Survival (%) after 1 and 5 years after lung cancer diagnosis stratified by database, sex and calendar year**

| **Calendar Year** | **Time (years)** | **% Survival (95% CI)** | **Sex** | **Database** |
| --- | --- | --- | --- | --- |
| 2000 to 2004 | 1 | 37.23 (36.29 - 38.20) | Both | Aurum |
| 2005 to 2009 | 1 | 35.17 (34.46 - 35.89) |  |  |
| 2010 to 2014 | 1 | 40.06 (39.40 - 40.72) |  |  |
| 2015 to 2019 | 1 | 48.02 (47.40 - 48.65) |  |  |
| 2000 to 2004 | 1 | 33.43 (32.14 - 34.77) |  | GOLD |
| 2005 to 2009 | 1 | 35.28 (34.36 - 36.21) |  |  |
| 2010 to 2014 | 1 | 39.58 (38.67 - 40.51) |  |  |
| 2015 to 2019 | 1 | 45.20 (44.13 - 46.30) |  |  |
| 2020 to 2021 | 1 | 44.12 (42.06 - 46.28) |  |  |
| 2000 to 2004 | 1 | 40.23 (38.72 - 41.80) | Female | Aurum |
| 2005 to 2009 | 1 | 37.95 (36.84 - 39.09) |  |  |
| 2010 to 2014 | 1 | 43.11 (42.14 - 44.10) |  |  |
| 2015 to 2019 | 1 | 52.41 (51.52 - 53.32) |  |  |
| 2000 to 2004 | 1 | 37.38 (35.32 - 39.56) |  | GOLD |
| 2005 to 2009 | 1 | 38.83 (37.43 - 40.27) |  |  |
| 2010 to 2014 | 1 | 42.64 (41.32 - 44.01) |  |  |
| 2015 to 2019 | 1 | 48.98 (47.49 - 50.53) |  |  |
| 2020 to 2021 | 1 | 46.26 (43.37 - 49.35) |  |  |
| 2000 to 2004 | 1 | 35.24 (34.04 - 36.48) | Male | Aurum |
| 2005 to 2009 | 1 | 33.14 (32.23 - 34.09) |  |  |
| 2010 to 2014 | 1 | 37.45 (36.57 - 38.34) |  |  |
| 2015 to 2019 | 1 | 44.07 (43.22 - 44.93) |  |  |
| 2000 to 2004 | 1 | 30.77 (29.14 - 32.48) |  | GOLD |
| 2005 to 2009 | 1 | 32.49 (31.30 - 33.73) |  |  |
| 2010 to 2014 | 1 | 36.82 (35.59 - 38.09) |  |  |
| 2015 to 2019 | 1 | 41.29 (39.78 - 42.85) |  |  |
| 2020 to 2021 | 1 | 41.99 (39.12 - 45.07) |  |  |
| 2000 to 2004 | 5 | 10.47 (9.24 - 11.85) | Both | Aurum |
| 2005 to 2009 | 5 | 9.09 (8.25 - 10.02) |  |  |
| 2010 to 2014 | 5 | 12.53 (11.56 - 13.58) |  |  |
| 2015 to 2019 | 5 | 19.51 (18.33 - 20.77) |  |  |
| 2000 to 2004 | 5 | 8.89 (7.30 - 10.82) |  | GOLD |
| 2005 to 2009 | 5 | 8.46 (7.00 - 10.22) |  |  |
| 2010 to 2014 | 5 | 12.96 (11.84 - 14.19) |  |  |
| 2015 to 2019 | 5 | 17.31 (15.81 - 18.94) |  |  |
| 2000 to 2004 | 5 | 12.91 (10.73 - 15.54) | Female | Aurum |
| 2005 to 2009 | 5 | 11.03 (9.70 - 12.54) |  |  |
| 2010 to 2014 | 5 | 15.12 (13.72 - 16.67) |  |  |
| 2015 to 2019 | 5 | 24.41 (22.84 - 26.09) |  |  |
| 2000 to 2004 | 5 | 10.39 (7.52 - 14.34) |  | GOLD |
| 2005 to 2009 | 5 | 8.90 (6.42 - 12.34) |  |  |
| 2010 to 2014 | 5 | 15.43 (13.68 - 17.40) |  |  |
| 2015 to 2019 | 5 | 21.06 (18.80 - 23.58) |  |  |
| 2000 to 2004 | 5 | 8.94 (7.56 - 10.57) | Male | Aurum |
| 2005 to 2009 | 5 | 7.69 (6.63 - 8.91) |  |  |
| 2010 to 2014 | 5 | 10.31 (9.02 - 11.79) |  |  |
| 2015 to 2019 | 5 | 15.05 (13.24 - 17.10) |  |  |
| 2000 to 2004 | 5 | 7.72 (6.06 - 9.85) |  | GOLD |
| 2005 to 2009 | 5 | 8.24 (6.95 - 9.76) |  |  |
| 2010 to 2014 | 5 | 10.69 (9.28 - 12.31) |  |  |
| 2015 to 2019 | 5 | 13.49 (11.64 - 15.64) |  |  |
